# Digital wellbeing in the social media era: dual pathway analysis of emotional and behavioural distress indicators and public health utility

**DOI:** 10.64898/2026.10.09.26365101

**Authors:** Suzana Younas, Tertsegha Joseph Anande

## Abstract

Social media language and digital lifestyle behaviours are promoted as scalable signals of population mental health, but predictive models are often evaluated in ways that overstate accuracy. We analysed 20,000 tweets from 72 users, labelled by whether the account belonged to a user identified as depressed, and two synthetic datasets (100,000 and 30,000 records) linking screen time, sleep and social media use with self-reported stress. Under a conventional tweet-level split, a term-weighted logistic regression appeared accurate (area under the receiver operating characteristic curve 0.835), but when whole users were held out its performance fell to near chance (0.595; 95% confidence interval 0.505–0.686), because the model largely recognised users and their topics. Depression-associated tweets were more often negative (31.7% versus 24.2%), yet sentiment alone could not identify unseen users (0.539). The two behavioural datasets gave opposite answers: strong discrimination in one (0.926), driven by short sleep, and chance in the other (0.506), so synthetic results reflect how the data were generated. At a population prevalence of 10%, positive predictive values would be 0.10–0.30. Digital distress signals should be validated across individuals, on real linked data and at realistic prevalence before informing public health surveillance or prevention.

## 1 Introduction

Mental disorders are among the ten leading causes of disability worldwide, and their burden has not fallen since 1990[1]. The World Health Organization estimated that nearly one billion people were living with a mental disorder in 2019 and has called for population-wide approaches that address modifiable determinants of mental health[2]. Chronic psychological stress is one such determinant: sustained activation of physiological stress systems contributes to cumulative “allostatic load” and increases the risk of depression, anxiety and cardiovascular disease[3]. Population mental health surveillance nevertheless still relies largely on periodic surveys, which are costly, slow and coarse in time and geography.

Digital footprints offer a potential complement. People increasingly express their emotional states on social media, and language markers such as negative affect, self-focus and references to sleep or bodily symptoms are associated with depression and distress[4–7]. Analysing such content for public health purposes, termed infoveillance, could provide near-real-time population signals[8]. In parallel, heavy screen use and short or disrupted sleep are associated with poorer mental health[9–12], and these behaviours are modifiable targets for public health guidance[13]. The size of these associations remains debated: large-sample analyses report small effects[14, 15], whereas quasi-experimental evidence suggests a causal contribution of social media to poorer mental health[16].

Many computational studies report high accuracy for detecting distress from either source[17–19]. Critical reviews, however, warn that three problems can inflate reported performance. Proxy labels may capture topic rather than mental state; evaluation designs may allow the same individual to appear in both training and test data; and models often fail to generalise beyond the dataset on which they were developed[20–23]. Openly shared datasets, including synthetic ones, are widely reused in applied and student work, often without these checks. Moreover, accuracy measured on balanced test sets says little about how many people would be wrongly flagged at the much lower prevalence found in the general population. For public health, the questions that matter are whether a signal works for people it has never seen, and what it would yield in a real population.

Multimodal studies that combine social media with sensing or survey data are promising[24–27], but linked datasets are scarce, and most studies examine a single pathway. Here we evaluated three widely reused open datasets within one reproducible framework: an emotional pathway based on Twitter language and sentiment, and a behavioural pathway based on screen time, sleep and social media use, analysed in two independently generated synthetic datasets, one of which was the dataset of the original project. We asked how well each pathway discriminates psychological distress, how strongly the evaluation design changes the answer, and what the results would imply if the models were deployed for population screening (Fig. 1).

**Fig 1.**
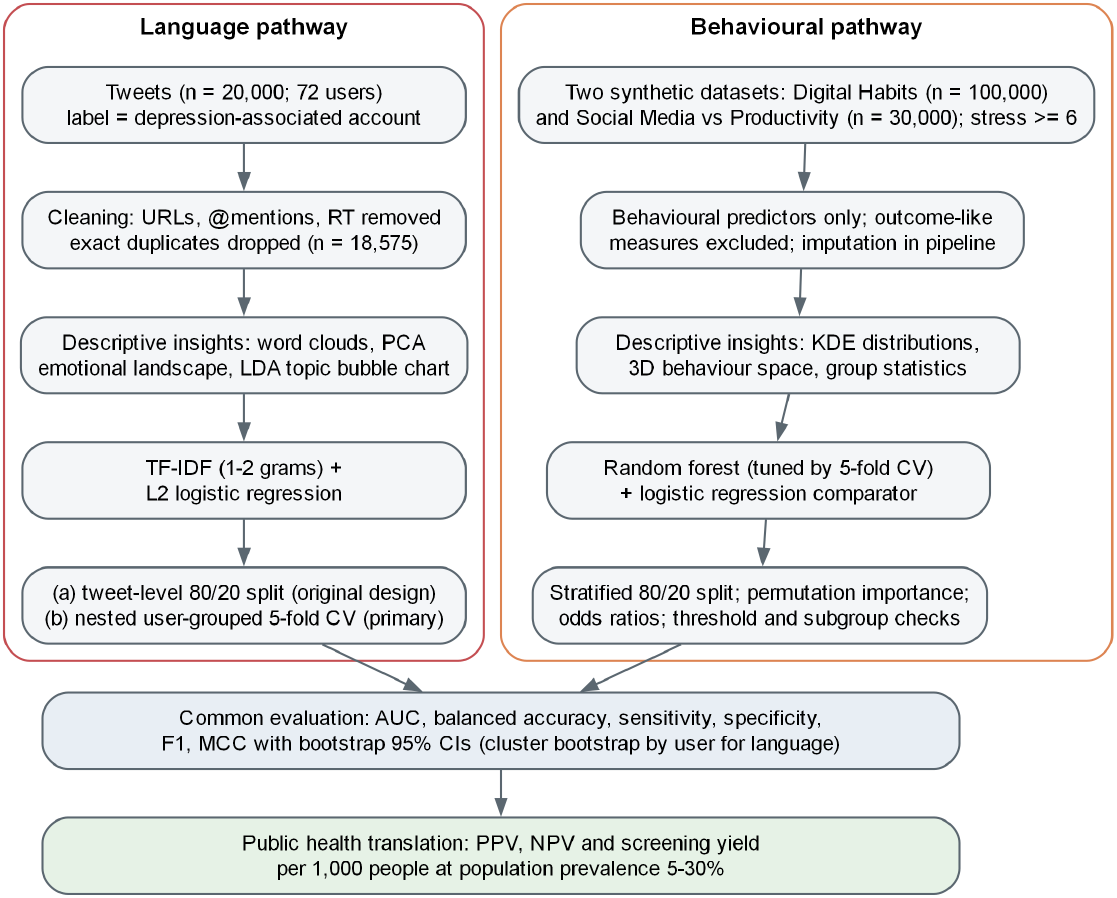
Analytical workflow. The language and behavioural pathways were processed, modelled and evaluated independently, then compared on common metrics and translated into population-level indicators.

## 2 Results

### 2.1 Behavioural distress markers in two synthetic datasets

The Digital Habits dataset comprised 100,000 synthetic records, of which 62,669 (62.7%) reported higher stress (score ≥6). Higher-stress records showed more screen time, less sleep and more TikTok use, whereas the number of platforms used did not differ between groups (Table 1). Sleep showed the largest standardised difference (−1.40). The density of sleep duration peaked at about 6.5 hours for higher-stress records, below the 7 hours recommended for adults[28], and at about 8 hours for lower-stress records; screen time peaked at about 6.5 and 5 hours, respectively (Fig. 2a,b). The distributions overlapped substantially, and sharp density changes at the range limits indicated truncation, consistent with simulated data. A three-dimensional view of screen time, sleep and TikTok use showed the same pattern of graded rather than separated groups (Supplementary Fig. S1).

**Table 1.** Behavioural features by stress group in the two synthetic datasets. Values are mean (SD).

| Feature | All | Higher stress | Lower stress | SMD |
| --- | --- | --- | --- | --- |
| *Digital Habits* ($N = 100,000$) | | | | |
| Screen time (hours/day) | 6.00 (1.99) | 6.60 (1.85) | 5.00 (1.79) | 0.88 |
| Sleep duration (hours/night) | 6.99 (1.47) | 6.36 (1.27) | 8.04 (1.13) | -1.40 |
| Hours on TikTok | 2.40 (1.08) | 2.71 (1.08) | 1.89 (0.86) | 0.84 |
| Social media platforms used | 3.00 (1.41) | 3.00 (1.42) | 2.99 (1.41) | 0.01 |
| Stress level (1–10 scale) | 6.18 (2.05) | 7.45 (1.29) | 4.04 (1.06) | – |
| *Social Media vs Productivity* ($N = 28,096$) | | | | |
| Daily social media time (hours) | 3.12 (2.07) | 3.11 (2.06) | 3.12 (2.09) | 0.00 |
| Sleep duration (hours/night) | 6.50 (1.47) | 6.50 (1.47) | 6.50 (1.46) | 0.00 |
| Screen time before sleep (hours) | 1.03 (0.65) | 1.02 (0.65) | 1.03 (0.65) | 0.00 |
| Notifications per day | 59.98 (7.75) | 59.94 (7.75) | 60.02 (7.74) | -0.01 |
| Stress level (1–10 scale) | 5.51 (2.87) | 8.00 (1.42) | 3.02 (1.41) | – |

Higher stress: score $\geq 6$ . Digital Habits: $n = 62,669$ higher and 37,331 lower. Social Media vs Productivity (records with a stress score): $n = 14,066$ higher and 14,030 lower; means use available values (7–9% missing for social media time, sleep and screen time before sleep). SMD, standardised mean difference (higher minus lower).

**Fig 2.**
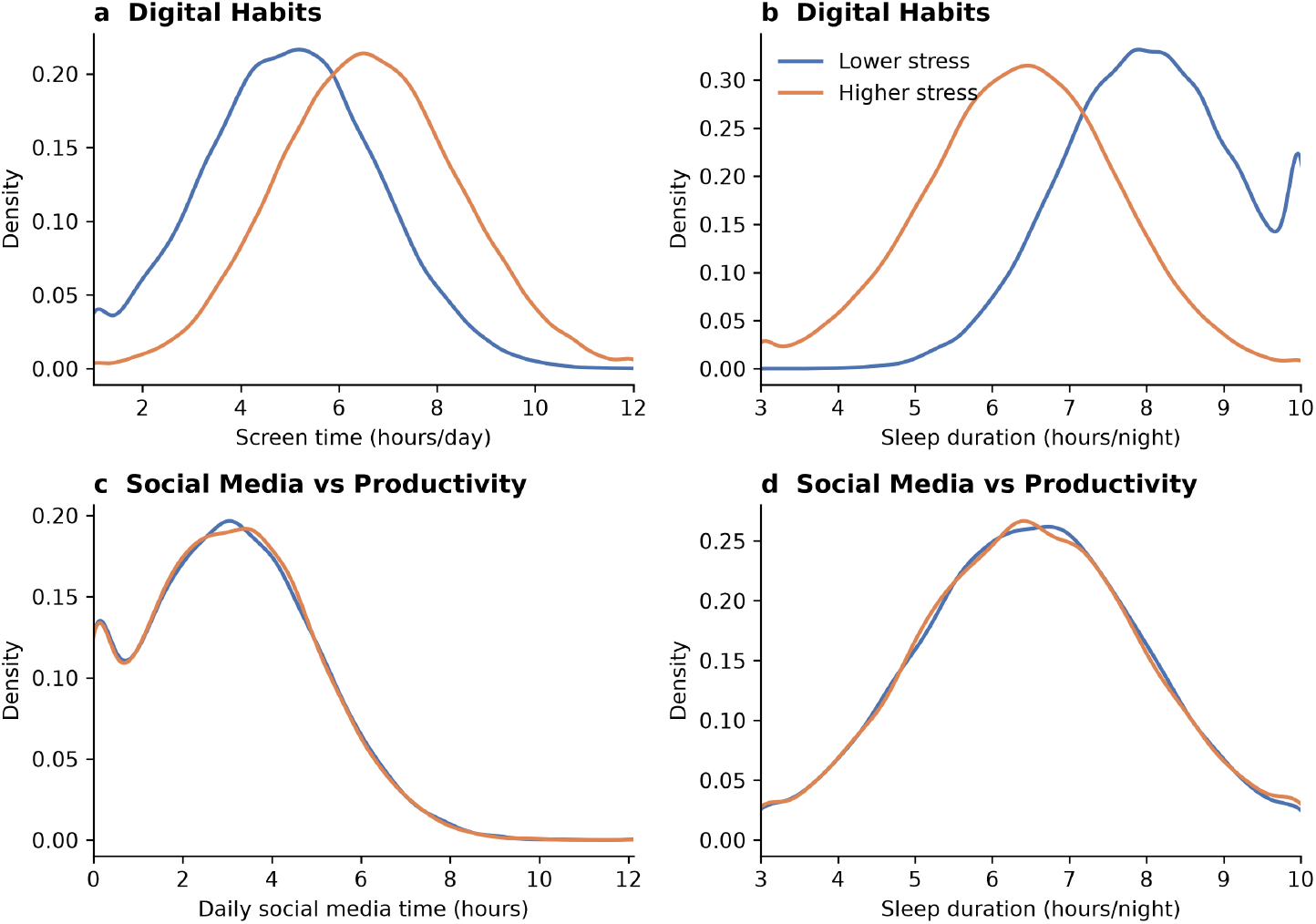
Distributions of behavioural markers by stress group in the two synthetic datasets. Kernel density estimates for records with higher (score ≥6) and lower self-reported stress: Digital Habits, (**a**) daily screen time and (**b**) nightly sleep duration; Social Media vs Productivity, (**c**) daily social media time and (**d**) nightly sleep duration.

On a held-out test set of 20,000 records, a tuned random forest achieved an area under the receiver operating characteristic curve (AUC) of 0.926 (95% CI 0.923–0.930), with a sensitivity of 0.893 and a specificity of 0.773 (Table 2). A plain logistic regression performed equivalently (AUC 0.927, 95% CI 0.924–0.931), so the more complex model added no benefit. Adding the mood score as a predictor did not change performance (AUC 0.926), and results were similar when stress was dichotomised at other thresholds (≥5: AUC 0.936; ≥7: AUC 0.928). Sleep duration dominated permutation importance (mean AUC decrease 0.283), followed by screen time (0.067) and Tik-Tok use (0.034); the number of platforms contributed nothing (0.000). In a mutually adjusted logistic model, each additional hour of sleep was associated with an odds ratio for higher stress of 0.17 (95% CI 0.166–0.174), each hour of screen time with 1.80 (1.77–1.83), each hour on TikTok with 2.40 (2.33–2.48) and each additional platform with 1.00 (0.99–1.02). Classification errors were concentrated near the threshold: 41% of records with a stress score of 5 and 28% with a score of 6 were misclassified, compared with fewer than 8% at scores of 3 or below or 7 or above (Supplementary Fig. S2b).

**Table 2.** Discrimination performance with 95% confidence intervals: (**a**) behavioural pathway in the two synthetic datasets; (**b**) emotional pathway.

| **a** | Digital Habits |  | Social Media vs Productivity |  |
| --- | --- | --- | --- | --- |
|  | Random forest | Logistic regression | Random forest | Logistic regression |
| Baseline accuracy | 0.627 | 0.627 | 0.501 | 0.501 |
| AUC | 0.926<br>(0.923–0.930) | 0.927<br>(0.924–0.931) | 0.506<br>(0.490–0.521) | 0.498<br>(0.482–0.513) |
| Accuracy | 0.848<br>(0.843–0.853) | 0.847<br>(0.842–0.852) | 0.506<br>(0.493–0.519) | 0.499<br>(0.486–0.512) |
| Balanced accuracy | 0.833<br>(0.827–0.838) | 0.833<br>(0.828–0.839) | 0.506<br>(0.493–0.519) | 0.499<br>(0.486–0.512) |
| Precision | 0.868<br>(0.862–0.874) | 0.871<br>(0.865–0.876) | 0.507<br>(0.488–0.525) | 0.500<br>(0.482–0.518) |
| Sensitivity | 0.893<br>(0.887–0.898) | 0.888<br>(0.883–0.894) | 0.503<br>(0.484–0.521) | 0.528<br>(0.510–0.547) |
| Specificity | 0.773<br>(0.763–0.782) | 0.778<br>(0.769–0.788) | 0.509<br>(0.490–0.527) | 0.470<br>(0.452–0.489) |
| F1 score | 0.880<br>(0.876–0.884) | 0.879<br>(0.875–0.883) | 0.505<br>(0.488–0.520) | 0.514<br>(0.498–0.529) |
| MCC | 0.672<br>(0.662–0.683) | 0.672<br>(0.661–0.682) | 0.012<br>(−0.014–0.037) | −0.002<br>(−0.028–0.025) |

| **b** | Emotional pathway (logistic regression) |  |
| --- | --- | --- |
|  | Tweet-level split | User-grouped CV |
| Baseline accuracy | 0.520 | 0.520 |
| AUC | 0.835<br>(0.822–0.848) | 0.595<br>(0.505–0.686) |
| Accuracy | 0.741<br>(0.727–0.755) | 0.557<br>(0.468–0.652) |
| Balanced accuracy | 0.738<br>(0.724–0.752) | 0.552<br>(0.485–0.628) |
| Precision | 0.721<br>(0.702–0.740) | 0.561<br>(0.362–0.800) |
| Sensitivity | 0.819<br>(0.802–0.836) | 0.677<br>(0.599–0.756) |
| Specificity | 0.657<br>(0.634–0.679) | 0.426<br>(0.318–0.556) |
| F1 score | 0.767<br>(0.752–0.781) | 0.614<br>(0.466–0.745) |
| MCC | 0.484<br>(0.455–0.512) | 0.107<br>(−0.031–0.254) |

Evaluated on held-out synthetic records (Digital Habits, 20,000; Social Media vs Productivity, 5,620), 3,715 held-out tweets (tweet-level split) and 18,575 tweets from 72 users (user-grouped cross-validation, CV). Positive class: higher stress (behavioural) and depression-associated account (emotional). Baseline accuracy is that of always predicting the majority class. User-grouped confidence intervals use a cluster bootstrap that resamples users. AUC, area under the receiver operating characteristic curve; MCC, Matthews correlation coefficient.

The Social Media vs Productivity dataset, the dataset of the original project, told the opposite story. Of its 30,000 records, 1,904 lacked a stress score; of the remaining 28,096, 14,066 (50.1%) reported higher stress. Self-reported stress was almost uniformly distributed across the 1–10 scale, and no behaviour differed between stress groups: standardised mean differences were at most 0.02 in absolute value, and the largest absolute correlation between stress and any other variable was 0.012 (Table 1; Fig. 2c,d). On a held-out test set of 5,620 records, the tuned random forest performed at chance (AUC 0.506, 95% CI 0.490–0.521; balanced accuracy 0.506), as did logistic regression (AUC 0.498, 95% CI 0.482–0.513) (Table 2). A model given every variable in the dataset, including the excluded productivity, burnout and job-satisfaction measures and the demographics, also performed at chance (AUC 0.497, 95% CI 0.481– 0.512), as did models using the original project’s threshold (stress ≥7: AUC 0.499) or a lower one (≥5: AUC 0.502). Mutually adjusted odds ratios were null for daily social media time (1.00, 95% CI 0.99–1.02), sleep (1.00, 0.98–1.02) and screen time before sleep (0.99, 0.95–1.03); one of ten behaviours reached nominal significance (digital-wellbeing features enabled: 0.94, 0.88–1.00; *p* = 0.043), as expected by chance across ten tests. Performance was equally at chance across genders (AUC 0.502–0.564) and all six job types (AUC 0.497–0.517; Supplementary Table S2).

### 2.2 Language and sentiment of depression-associated tweets

After cleaning and de-duplication, the corpus comprised 18,575 tweets (9,662 depression-associated; 8,913 comparison) from 72 users, who contributed a median of 114 tweets each (range 1–2,055). “Depression” was the most frequent word in depression-associated tweets, and health and mental-health vocabulary featured prominently, including “overcome”, “help”, “health”, “mental”, “anxiety”, “sleep”, “pain”, “addiction”, “treatment”, “therapy” and “suicide” (Fig. 3a). However, every-day words such as “like”, “love”, “people”, “good” and “day” were the next most frequent after “depression”, showing that most tweets from these accounts concerned ordinary life rather than distress. Comparison tweets shared the same everyday vocabulary but featured politics and news (“trump”, “president”, “russia”, “obama”, “vote”) and social interaction (“thank”, “thanks”, “hello”, “follow”), with little health-related language (Fig. 3b). The two classes therefore differed in topic as well as in emotional content.

**Fig 3.**
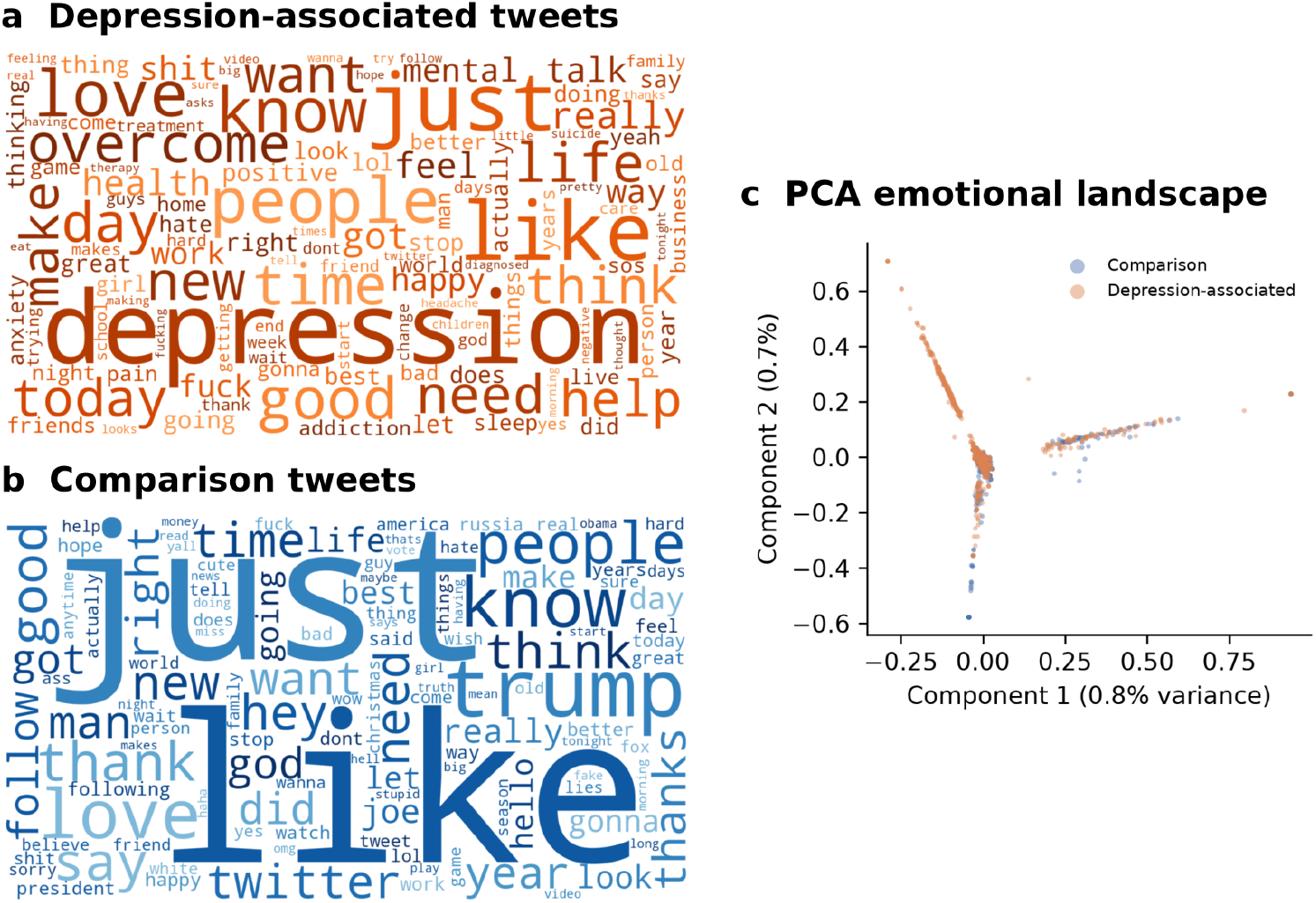
Language of depression-associated and comparison tweets. Word clouds of the most frequent words in (**a**) depression-associated and (**b**) comparison tweets, restricted to words written by at least five different users so that account-specific terms cannot identify individuals. (**c**) Projection of term-weighted tweet vectors onto the first two principal components (random sample of 5,000 tweets).

The first two principal components of the term-weighted text vectors explained only 1.5% of the variance, as is typical of sparse text (Fig. 3c). Most tweets (89.1%) fell in a dense central cluster in which the two classes were evenly mixed (50.4% depression-associated). One arm, defined by “depression”, “overcome”, “treatment”, “therapy” and “health”, contained 754 tweets (4.1%), of which 98.8% were depression-associated, written by 58 different users. A second arm, defined by everyday affective words (“like”, “love”, “hate”), was mixed (52.7%). Explicit mental-health language was thus a distinctive and widely shared marker, but it appeared in only a small fraction of tweets. A six-topic latent Dirichlet allocation model told the same story: the share of depression-associated tweets ranged from 42.6% in a politics-related topic to 68.3% in a topic defined by “depression”, “overcome” and “thanks” (corpus average 52.0%), and every topic contained both classes (Supplementary Fig. S3).

Depression-associated tweets were more often negative than comparison tweets (31.7% vs 24.2%) and less often neutral (29.2% vs 37.4%), whereas positive tweets were equally common (39.1% vs 38.4%) (Fig. 4a). Expressed distress therefore appeared as additional negative affect rather than reduced positive affect. At the user level, depression-associated users showed a moderately higher intensity of negative affect (mean negative score 0.128 vs 0.081; Cohen’s *d* = 0.56; *p* = 0.037). Their mean compound sentiment was lower (0.020 vs 0.140; *d* = −0.40), but this difference was not significant (*p* = 0.51). It was driven by eight depression-associated users whose average sentiment was strongly negative, seven of whom posted only one to four tweets (Fig. 4b). The group medians were similar (0.086 vs 0.099), and without these eight users the depression-associated mean was 0.134. In mixed models that used all tweets while accounting for clustering within users (intraclass correlation 0.08–0.11), the difference in negative affect was in the same direction but not statistically significant (*β* = 0.023, 95% CI −0.003 to 0.049; *p* = 0.078; Supplementary Table S1). A classifier based on sentiment alone performed at chance on unseen users (AUC 0.539, 95% CI 0.489–0.600; balanced accuracy 0.546).

**Fig 4.**
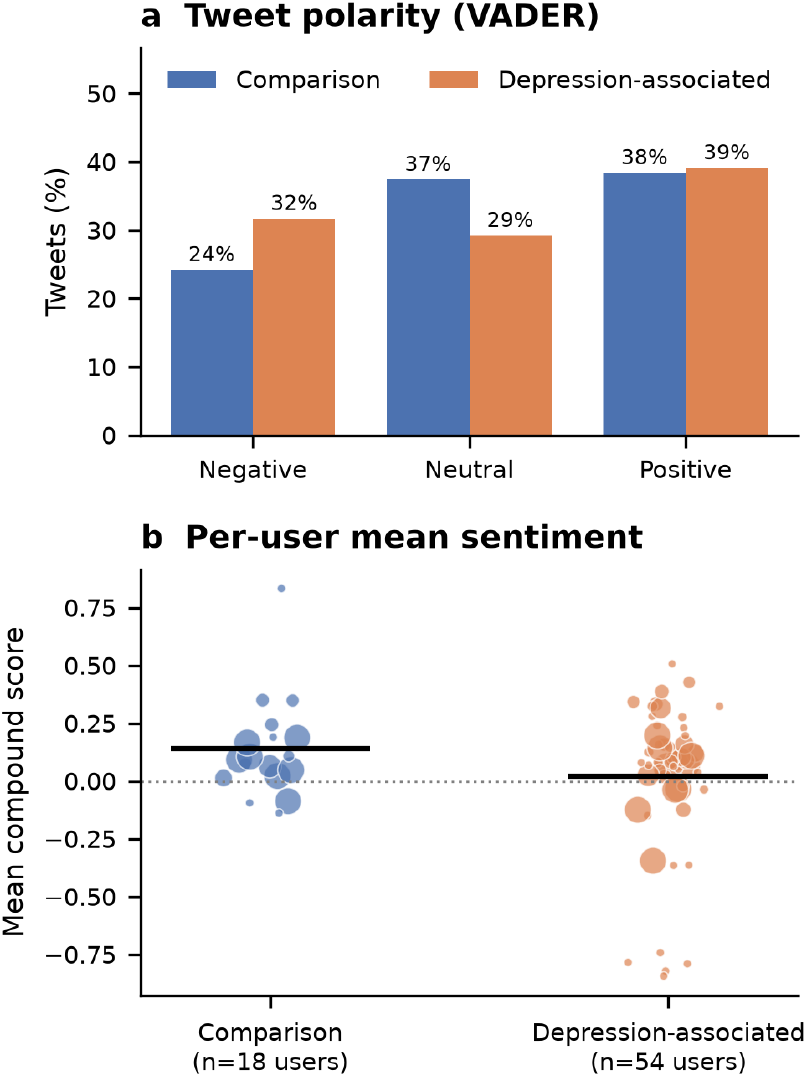
Sentiment of depression-associated and comparison tweets (VADER). (**a**) Share of negative, neutral and positive tweets in each class. (**b**) Mean compound sentiment score per user; bubble size is proportional to the number of tweets, and black lines show group means.

### 2.3 Evaluation design determines apparent language-model accuracy

With the conventional tweet-level split, the language model appeared to discriminate well (AUC 0.835, 95% CI 0.822–0.848; Table 2). When whole users were held out, the AUC fell to 0.595 (95% CI 0.505–0.686) and the balanced accuracy to 0.552 (95% CI 0.485–0.628); both intervals include or approach chance (Fig. 5). The same figure shows the two behavioural datasets: near-perfect discrimination in one and chance in the other. AUCs in the individual outer folds ranged from 0.53 to 0.70. On unseen users, the model labelled most tweets as depression-associated, identifying 68% of depression-associated tweets but only 43% of comparison tweets (Supplementary Fig. S2a). Averaging tweet probabilities per user classified 50 of 54 depression-associated users but only 6 of 18 comparison users correctly (user-level AUC 0.818), an imprecise estimate given only 72 users. The terms most strongly weighted towards the depression-associated class included mental and physical health vocabulary, but also account-specific hashtags and fan-community terms, whereas terms weighted towards the comparison class were dominated by politics, news and promotional hashtags. The model therefore relied partly on who was writing and which topics they followed, which explains its failure on new users. An earlier version of this analysis that did not remove user handles or duplicate tweets reported an accuracy of 0.841; removing them reduced tweet-level accuracy to 0.741 even before users were held out.

**Fig 5.**
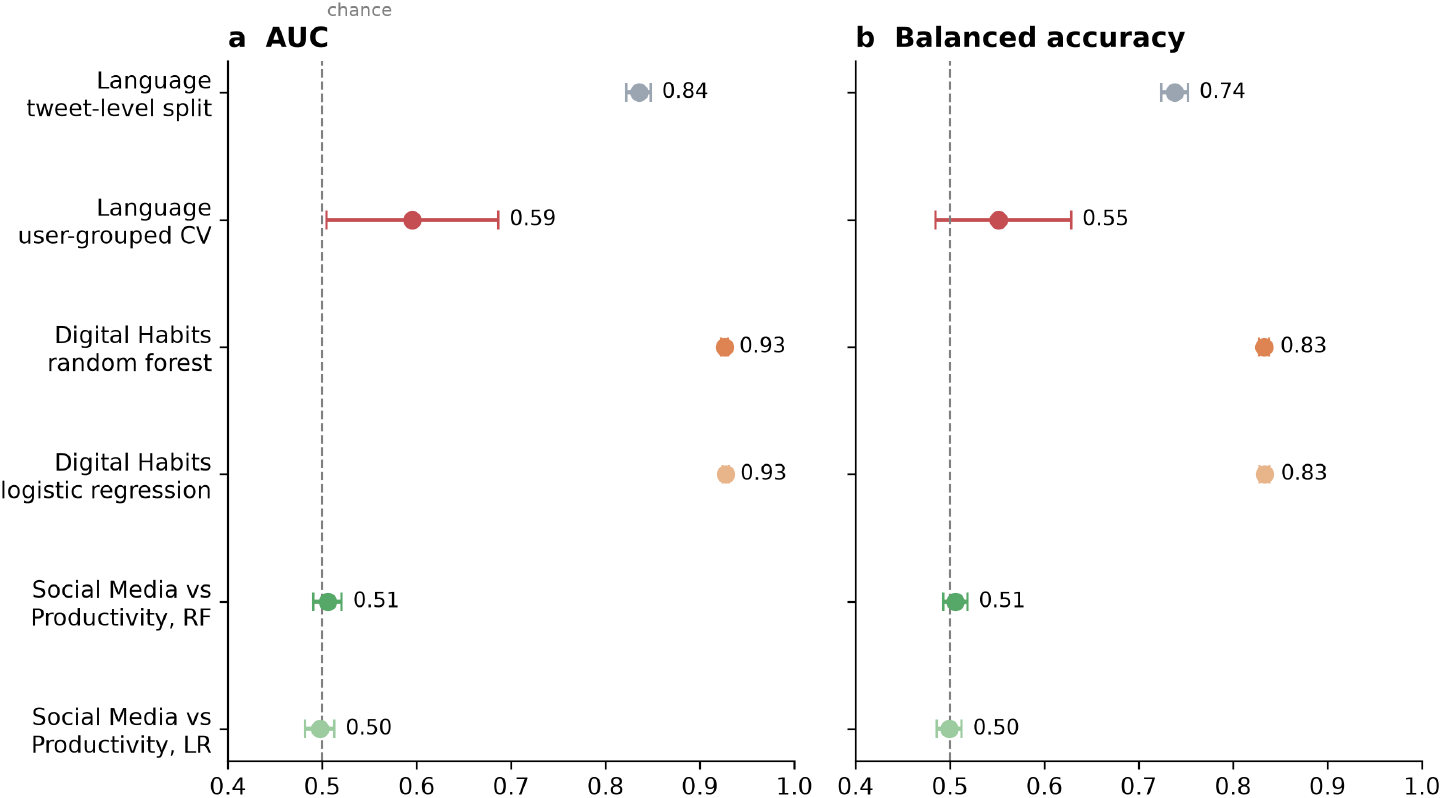
Effect of evaluation design on discrimination. (**a**) Area under the receiver operating characteristic curve and (**b**) balanced accuracy, with 95% confidence intervals, for the emotional pathway under a tweet-level split and under user-grouped cross-validation, and for the random forest (RF) and logistic regression (LR) models on held-out records of the two synthetic behavioural datasets. The dashed line marks chance performance.

### 2.4 Calibration

Calibration told the same story as discrimination (Supplementary Fig. S4). On unseen users, the language model’s predicted probabilities were far too extreme: the calibration slope was 0.32 (95% CI 0.08–0.58), and its Brier score of 0.261 was no better than assigning every tweet the overall prevalence (scaled Brier −0.05; 95% CI −0.35 to 0.06). Under the tweet-level split, the same model was miscalibrated in the opposite direction, with predictions too cautious (slope 1.56; Brier 0.168). The Digital Habits models were almost perfectly calibrated (random forest slope 0.99 (95% CI 0.96–1.02), calibration-in-the-large 0.02, Brier 0.106; logistic regression slope 0.97), as expected for data generated by a simple rule. The Social Media vs Productivity models carried no information: their calibration slopes did not differ from zero (random forest 0.06 (95% CI −0.22 to 0.33); logistic regression −0.39 (95% CI −1.81 to 0.94)), and their Brier scores (0.252 and 0.250) equalled that of predicting the prevalence for everyone.

### 2.5 Implications for population screening

Applying each primary model’s sensitivity and specificity to realistic prevalences showed that neither would be suitable for flagging individuals (Table 3). At 10% prevalence, only 12% of people flagged by the emotional model and 30% of those flagged by the behavioural model would truly be positive. Per 1,000 people screened, the behavioural model would identify about 89 true cases at the cost of 204 false alarms, and the emotional model 68 true cases at the cost of 516 false alarms. For the Social Media vs Productivity models, the positive predictive value simply equalled prevalence (0.10 at 10%), which is no better than selecting people at random.

**Table 3.**
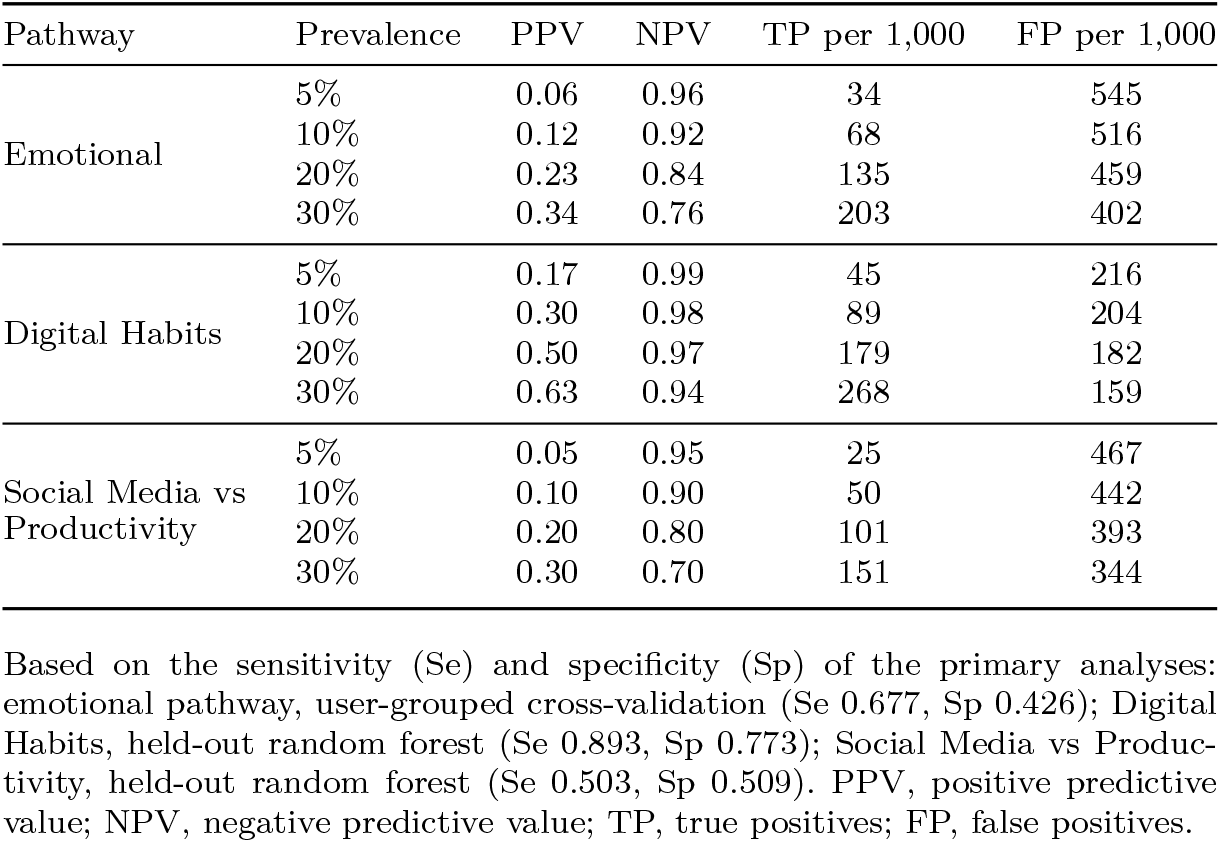
Expected predictive values and screening yield per 1,000 people at population prevalence.

| Pathway | Prevalence | PPV | NPV | TP per 1,000 | FP per 1,000 |
| --- | --- | --- | --- | --- | --- |
| Emotional | 5% | 0.06 | 0.96 | 34 | 545 |
|  | 10% | 0.12 | 0.92 | 68 | 516 |
|  | 20% | 0.23 | 0.84 | 135 | 459 |
|  | 30% | 0.34 | 0.76 | 203 | 402 |
| Digital Habits | 5% | 0.17 | 0.99 | 45 | 216 |
|  | 10% | 0.30 | 0.98 | 89 | 204 |
|  | 20% | 0.50 | 0.97 | 179 | 182 |
|  | 30% | 0.63 | 0.94 | 268 | 159 |
| Social Media vs Productivity | 5% | 0.05 | 0.95 | 25 | 467 |
|  | 10% | 0.10 | 0.90 | 50 | 442 |
|  | 20% | 0.20 | 0.80 | 101 | 393 |
|  | 30% | 0.30 | 0.70 | 151 | 344 |

Based on the sensitivity (Se) and specificity (Sp) of the primary analyses: emotional pathway, user-grouped cross-validation (Se 0.677, Sp 0.426); Digital Habits, held-out random forest (Se 0.893, Sp 0.773); Social Media vs Productivity, held-out random forest (Se 0.503, Sp 0.509). PPV, positive predictive value; NPV, negative predictive value; TP, true positives; FP, false positives.

## 3 Discussion

This study examined how well social media language and digital lifestyle behaviours discriminate psychological distress, and how robust that performance is to the evaluation design. A language model that appeared accurate under a tweet-level split performed close to chance on users it had not seen. Its apparent skill came largely from recognising the writing, topics and communities of individual users rather than markers of distress that transfer to new people. This is consistent with evidence that social media mental health models generalise poorly across users, datasets and time[22, 23], and that proxy labels capture correlated topics rather than clinical states[20, 21]. Two features of the corpus made it especially vulnerable: only 72 accounts contributed all the tweets, and the comparison accounts differed systematically in topic. Health-related terms did carry signal, in line with psycholinguistic studies[29, 30], but they appeared in only a small fraction of tweets.

The sentiment analysis tells the same story. Depression-associated users expressed negative affect more often, consistent with the established association between depression and negative emotional language[5, 7], but their positive affect was not reduced, the difference was modest once clustering within users was taken into account, and sentiment alone performed at chance on new users. Differences that appear clear in pooled tweets can therefore shrink substantially when analysed at the level of the individual, which is the level that matters for public health use.

The behavioural pathway gave opposite answers in the two synthetic datasets. In the Digital Habits dataset, less sleep and more screen time accompanied higher stress, in the direction reported by systematic reviews[9–11, 31]. The magnitudes, however, are implausible for real data. A standardised difference of 1.4 in sleep and an odds ratio of 0.17 per hour are far larger than population-based estimates, in which digital technology use explains well under 1% of the variance in adolescent well-being[14]. Synthetic datasets reproduce the assumptions of their generator, and models trained on them mainly recover those assumptions; the equivalent performance of random forest and logistic regression further suggests a simple, largely linear generating process. The Social Media vs Productivity dataset, analysed with the same pipeline, showed the reverse problem: stress was generated almost independently of every behaviour, so no model, however flexible or however many variables it was given, could predict it. Two synthetic datasets built around the same question therefore produced near-perfect and chance-level discrimination. Neither result says anything about human behaviour; each reflects how its data were generated. Researchers who develop or teach with such datasets should treat model performance on them as a property of the simulation rather than as evidence, and should report the provenance of any dataset they use.

These findings have four implications for public health. First, a model intended for surveillance or prevention must work for people it has never seen. Evaluations that allow the same individuals in training and test data are common but, as shown here, can substantially overstate performance; agencies and funders should require user-level and external validation, reported against TRIPOD+AI[32], before relying on such tools. Second, digital signals are better suited to surveillance than to individual screening. Even the optimistic behavioural model would produce about two false alarms for every true case at 10% prevalence, and labelling individuals on this basis risks stigma, unnecessary contact with services and loss of trust. Classification errors matter much less when validated signals are aggregated over regions, age groups or time periods and analysed as trends, where they could complement slow periodic surveys and help detect emerging population distress[8, 33]. Third, sleep and screen time, unlike language, are modifiable. Real-world evidence, although of smaller effect than in the synthetic data, supports guidance that adults sleep at least 7 hours per night[28] and that digital-wellbeing initiatives protect sleep, for example through device-free bedrooms and reduced night-time use[9, 13]. The absence of any association with the number of platforms, alongside clear associations with time-based measures, is a hypothesis worth testing in real cohorts, because it would favour messages about how long and when devices are used over which platforms are used; causal claims would require longitudinal or experimental evidence[16]. Fourth, people who post publicly about their health are not representative of the population, and models built on a few dozen English-speaking accounts cannot be assumed to work for other languages, cultures, age groups or platforms, so groups with low digital participation could be missed[2]. Open datasets, including synthetic ones, should document how they were constructed and labelled so that they are not mistaken for evidence about real populations[34].

This study has limitations. Both behavioural datasets are synthetic, so their associations reflect their generating processes. The emotional labels are undocumented account-level proxies for depression rather than validated measures of stress. The Twitter corpus contains only 72 users, so user-level estimates are imprecise, and its tweets (2009–2017) come from a single platform whose data access and user base have since changed. The two pathways involve different people, so multimodal fusion and within-person comparison were not possible. The design is cross-sectional. Demographics were available only in the Social Media vs Productivity dataset, where performance was uniformly at chance, so fairness could not be meaningfully assessed for a model with real discriminative ability. VADER is a general-purpose sentiment lexicon that does not capture specific emotions or sarcasm. Finally, we used a bag-of-words model; contextual language models may extract more transferable signal, but they are equally vulnerable to user leakage and must be evaluated in the same way. Future studies should use real, linked datasets in which the same consenting individuals provide social media text, sensed or self-reported behaviour and validated outcomes such as the Perceived Stress Scale[35], evaluate larger corpora with documented labelling across users, platforms and time, test multimodal fusion, audit fairness across subgroups, and co-design acceptable, privacy-preserving uses with public health practitioners and people with lived experience.

In conclusion, digital traces of language and lifestyle are often promoted as scalable tools for monitoring population mental health. Here, a language model that looked accurate largely failed on new users, sentiment differences were modest at the level of individuals, and two synthetic behavioural datasets built around the same question gave near-perfect and chance-level discrimination, showing that such results reflect how the data were generated. At realistic prevalences, both models would mislabel more people than they correctly identify. Before such signals inform public health practice, they need validation across individuals, real and linked data, and evaluation at the prevalence of the intended population.

## 4 Methods

### 4.1 Study design and reporting

This was a cross-sectional secondary analysis of three publicly available, de-identified datasets, analysed in two separate pathways because they contain different individuals and cannot be linked. Reporting follows the TRIPOD+AI guideline for prediction models that use machine learning[32]; the completed checklist is provided as Supplementary Information. No study protocol was prepared in advance, the study was not registered, and there was no patient or public involvement.

### 4.2 Data sources and outcomes

The emotional pathway used the *Depression: Twitter Dataset + Feature Extraction* corpus (version 2)[36], which contains 20,000 English tweets posted between August 2009 and January 2017 by 72 users and collected through the Twitter API. Each tweet carries its author’s account label: 54 accounts were labelled as belonging to depressed users (10,000 tweets) and 18 as non-depressed (10,000 tweets). The dataset creators do not document how the labels were assigned, so the outcome is a proxy for depression at the account level, which we call depression-associated distress rather than stress[21].

The behavioural pathway used the *Digital Habits vs. Mental Health* dataset (version 2)[37], which its creator describes as synthetic, that is, generated to simulate the behaviour of Generation Z users. It contains 100,000 records of daily screen time, sleep duration, TikTok use, number of social media platforms used (1–5), self-reported stress (1–10) and mood (1–10). There were no missing values, and the 1,865 exact duplicate rows were retained, as expected in simulated data with coarse values. Stress scores ≥6 were classified as higher stress; thresholds of ≥5 and ≥7 were examined in a sensitivity analysis. Mood was excluded from the predictors because it measures a closely related affective state (*r* = −0.64 with stress) and would risk outcome leakage.

The second behavioural dataset was the dataset of the original project: the *Social Media vs Productivity* dataset (version 1)[38], which its creator describes as simulating the behaviour of 30,000 people. It records daily social media time, sleep duration, screen time before sleep, number of notifications, preferred platform, work hours, breaks during work, coffee consumption, weekly offline hours, use of focus apps and digital-wellbeing features, self-reported stress (1–10), productivity, burnout and job satisfaction, and demographics (age, gender and job type). The 1,904 records without a stress score were excluded, leaving 28,096. Stress was dichotomised at ≥6, as in the Digital Habits dataset, with the original project’s threshold (≥7) and ≥5 as sensitivity analyses. Predictors were restricted to the digital and lifestyle behaviours; perceived and actual productivity, burnout days and job satisfaction were excluded as outcome-adjacent measures, as mood was for the Digital Habits dataset, and demographics were reserved for a subgroup analysis by gender and job type. Missing predictor values (7– 9% for social media time, sleep and screen time before sleep) were imputed with the training-set median, or the most frequent category, inside the model pipeline. To test whether any signal was being missed, a further model used every variable, including the excluded measures and demographics.

### 4.3 Text preprocessing and descriptive analyses

Tweets were lower-cased, and URLs, user mentions, the retweet marker “RT”, HTML entities, digits and punctuation were removed. Removing 1,425 exact duplicates left 18,575 tweets. Term frequency–inverse document frequency (TF–IDF) features[39] were computed over unigrams and bigrams (minimum document frequency 3, at most 50,000 features, sublinear term frequency, English stop words removed) and fitted on training data only. Descriptive analyses used only words written by at least five different users (2,307 words), so that account-specific hashtags, names or handles could neither identify individuals nor dominate the results. Word clouds showed the most frequent words in each class; a principal component analysis (PCA) of the TF–IDF vectors projected tweets onto two dimensions; and a latent Dirichlet allocation model with six topics assigned each tweet to its dominant topic.

### 4.4 Sentiment analysis

Sentiment was scored with VADER[40], a lexicon- and rule-based model designed for social media text, applied to lightly cleaned tweets (URLs, user mentions and “RT” removed) that retain the capitalisation, punctuation and emoticons VADER uses. Each tweet received negative, neutral and positive proportions and a normalised compound score (−1 to +1), and was classed as negative (compound ≤ −0.05), positive (≥0.05) or neutral. Because tweets are clustered within users, group differences were tested on per-user mean scores (Mann–Whitney U test and Cohen’s *d*) and at the tweet level with linear mixed models that included a random intercept for each user. A sentiment-only logistic regression (four VADER scores) was evaluated with the same nested user-grouped cross-validation as the TF–IDF model.

### 4.5 Models

The emotional pathway used L2-regularised logistic regression,

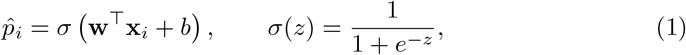

where **x**_*i*_ is the TF–IDF vector of tweet *i*, **w** the weight vector and *b* the intercept. Parameters minimise the penalised binary cross-entropy

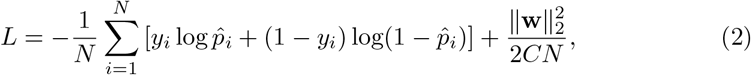

where *N* is the number of training samples, *y*_*i*_ ∈ {0, 1} the label and *C* ∈ { 0.01, 0.1, 1, 10} the inverse regularisation strength, selected by cross-validated AUC.

The behavioural pathway used a random forest[41] of *K* trees *h*_*k*_, with pre-dicted probability 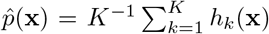 thresholded at 0.5. The number of trees ({100, 300, 500}), maximum depth ({none, 10, 20}) and minimum samples per split ({2, 10}) were tuned by stratified 5-fold cross-validated grid search on the training set; the selected model had 500 trees, a maximum depth of 10 and a minimum of 10 samples per split. Predictors were standardised using training-set statistics only. An unpenalised logistic regression on the same predictors served as a comparator. Predictor influence was assessed by permutation importance on the test set (mean decrease in AUC over 20 permutations), which is less biased than impurity-based importance[42], and mutually adjusted odds ratios per unit of each predictor were estimated on the full dataset.

### 4.6 Evaluation

For the emotional pathway, two designs were compared. In the tweet-level split, tweets were randomly divided into stratified training (80%) and test (20%) sets, replicating common practice in which the same users appear in both. In the primary analysis, nested user-grouped cross-validation, five outer folds held out whole users and *C* was tuned by an inner user-grouped 5-fold cross-validation on the training users; out-of-fold predictions were pooled for evaluation. Because tweets from the same user are not independent, 95% confidence intervals for this design were obtained from a cluster bootstrap that resampled users (5,000 replicates). User-level classification was assessed by averaging each user’s out-of-fold probabilities. For both behavioural datasets, a stratified 80/20 split was used, with the same random forest tuning and logistic regression comparator, and 95% confidence intervals from 5,000 bootstrap resamples of the test set. We report AUC, accuracy, balanced accuracy, sensitivity, specificity, precision, negative predictive value, F1 score and the Matthews correlation coefficient, together with the majority-class baseline. The pathways differ in population, outcome and class balance, so they are reported side by side but not compared statistically. Calibration was assessed for every primary model with the Brier score, the scaled Brier score (one minus the ratio of the Brier score to that of a model predicting the overall prevalence), calibration-in-the-large (the intercept of a logistic model with the logit of the predicted probability as an offset) and the calibration slope, with 1,000 bootstrap resamples for 95% confidence intervals (resampling users for the user-grouped language model), and with decile calibration plots. Each model was refitted with the same data, splits, random seeds and selected hyperparameters as in the main analysis and reproduced the reported AUC exactly.

### 4.7 Translation to population metrics

Predictive values depend on prevalence, so each primary model’s sensitivity (Se) and specificity (Sp) were applied to population prevalences *π* of 5%, 10%, 20% and 30%[1, 2]:

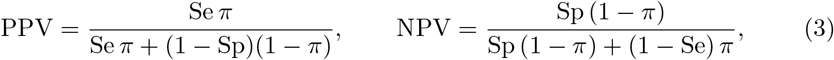

and the expected numbers of true and false positives per 1,000 people screened were calculated.

### 4.8 Software and reproducibility

Analyses used Python 3.11 with scikit-learn 1.9[43], pandas 3.0, statsmodels 0.15 and vaderSentiment, with a fixed random seed of 42. The complete analysis was run twice and reproduced identically, and every number reported in this article was checked automatically against the analysis outputs.

### 4.9 Use of large language models

The authors used Claude (Anthropic) to assist with reference verification, analysis code, figure generation and language editing. The authors reviewed and edited all output and take full responsibility for the content of this article.

### 4.10 Ethics

Ethical approval was not required because only publicly available, anonymised secondary data were used. No individual was contacted or re-identified, no tweet text is quoted, and descriptive analyses use only words written by at least five users, so that users cannot be traced through search[34]. All analyses were carried out in accordance with relevant guidelines and regulations.

## Data Availability

All three datasets analysed in this study are publicly available from Kaggle: Depression: Twitter Dataset + Feature Extraction (https://www.kaggle.com/datasets/infamouscoder/mental-health-social-media), Digital Habits vs. Mental Health (https://www.kaggle.com/datasets/abhishekdave9/digital-habits-vs-mental-health-dataset) and Social Media vs Productivity (https://www.kaggle.com/datasets/mahdimashayekhi/social-media-vs-productivity). The original project code is available at https://github.com/suzanayounas/Mental-Health-Vs-Social-Media, and the code that produces every result reported in the manuscript is available at https://github.com/Tertsegha1/digital-wellbeing-distress-signals.

https://www.kaggle.com/datasets/abhishekdave9/digital-habits-vs-mental-health-dataset

https://www.kaggle.com/datasets/mahdimashayekhi/social-media-vs-productivity

https://github.com/suzanayounas/Mental-Health-Vs-Social-Media

https://github.com/Tertsegha1/digital-wellbeing-distress-signals

## Data availability

All three datasets analysed in this study are publicly available from Kaggle: the *Depression: Twitter Dataset + Feature Extraction* corpus[36], the *Digital Habits vs. Mental Health* dataset[37] and the *Social Media vs Productivity* dataset[38].

## Code availability

The original project code, in which the dual-pathway analysis was conceived and first developed, is available at https://github.com/suzanayounas/Mental-Health-Vs-Social-Media. The code that produces every result, table and figure reported in this article, which builds on the original project with user-grouped validation, sentiment analysis, bootstrap confidence intervals and population-level translation, is available at https://github.com/Tertsegha1/digital-wellbeing-distress-signals.

## Supplementary information

Supplementary Figs. S1–S4 and Supplementary Tables S1 and S2 are provided in a separate Supplementary Information file, and the completed TRIPOD+AI checklist in a separate reporting-checklist file.

## Author contributions

S.Y. conceived the study and developed the original dual-pathway analysis: conceptualisation, methodology, software (original project code), data curation, formal analysis, visualisation, writing – original draft. T.J.A. contributed the methodological enhancements reported here: methodology (methodological enhancements: user-grouped validation, sentiment analysis, uncertainty quantification and public health translation), software (enhanced analysis code), formal analysis, validation, supervision, writing – review and editing. Both authors reviewed and approved the final manuscript.

