## Supplementary Information (Supplementary Figures S1-S4 and Tables S1-S2) for "Digital wellbeing in the social media era: dual pathway analysis of emotional and behavioural distress indicators and public health utility"

Suzana Younas and Tertsegha Joseph Anande
Department of Computing, Ulster University, Birmingham, United Kingdom

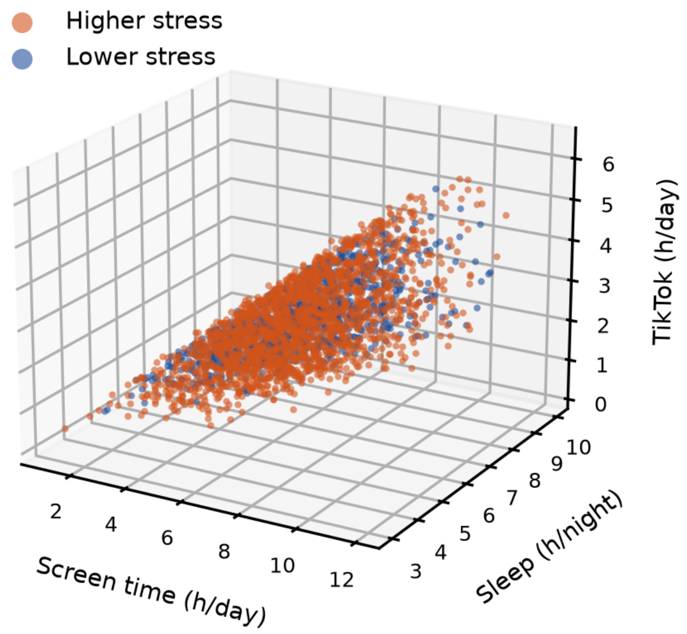


**Figure S1: Three-dimensional behaviour space.** Screen time, sleep duration and TikTok use for a random sample of 3,000 records from the synthetic behavioural dataset, coloured by stress group (higher stress: score ≥6). Screen time and TikTok use are strongly correlated ($r = 0.74$), so records lie along a diagonal band; higher-stress records are concentrated towards less sleep and more screen and TikTok use, with substantial overlap between groups.

**Table S1: User-level sentiment comparison and mixed-model estimates (VADER).**

| VADER score | Depression (54 users) | Comparison (18 users) | $d$ | $p$ (MWU) | Mixed model $\beta$ (95% CI); $p$ |
|---|---|---|---|---|---|
| Compound | 0.020 (0.317) | 0.140 (0.220) | −0.40 | 0.51 | −0.009 (−0.097, 0.079); 0.84 |
| Negative | 0.128 (0.093) | 0.081 (0.041) | 0.56 | 0.037 | 0.023 (−0.003, 0.049); 0.078 |
| Positive | 0.147 (0.073) | 0.146 (0.064) | 0.01 | 0.66 | 0.021 (−0.010, 0.052); 0.18 |
| % negative tweets | 35.6 (26.6) | 23.6 (14.1) | 0.50 | 0.17 | n/a |

Values are per-user means (SD). $d$: Cohen's $d$. MWU: Mann–Whitney U test. Mixed model: tweet-level linear mixed model with a random intercept per user; $\beta$ is the difference for depression-associated users.

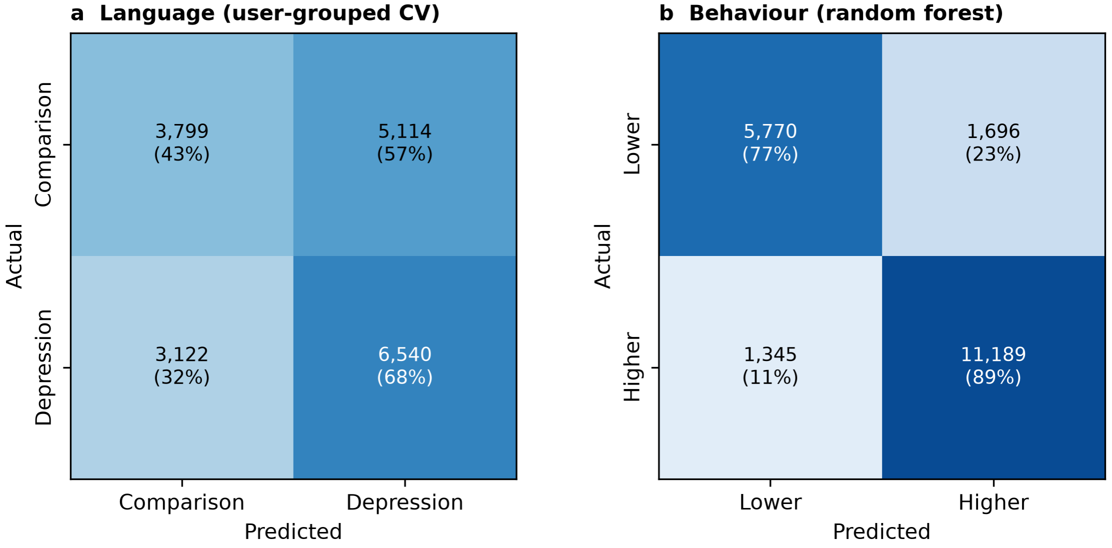


**Figure S2: Confusion matrices for the primary analyses.** (**a**) Emotional pathway: pooled out-of-fold predictions from nested user-grouped cross-validation (18,575 tweets from 72 users). (**b**) Behavioural pathway: random forest on the held-out test set of 20,000 synthetic records. Percentages are row percentages.

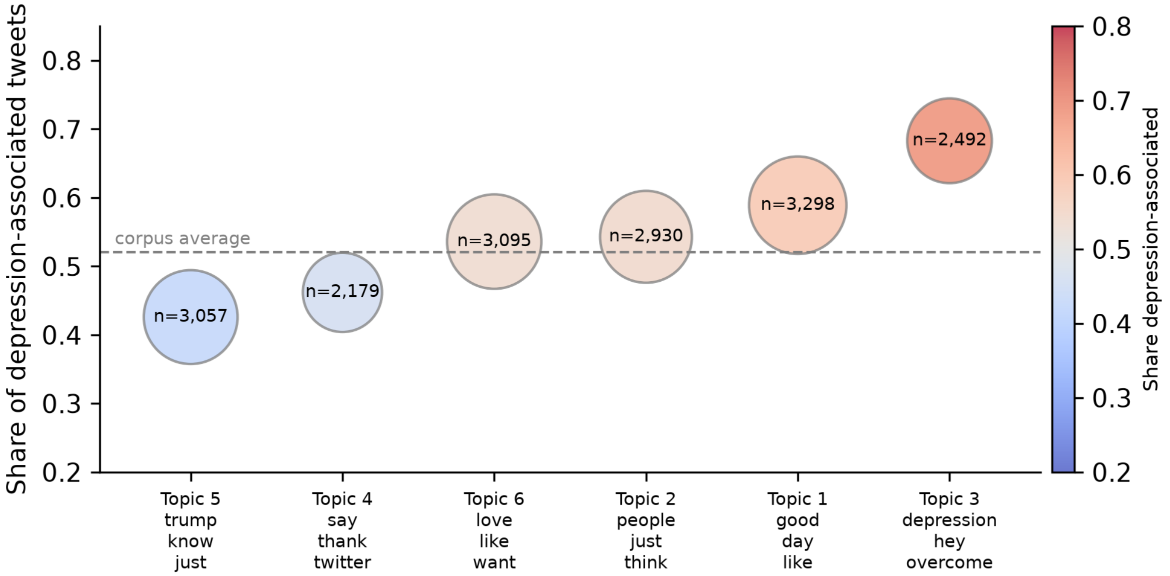


**Figure S3: Latent Dirichlet allocation topics.** Share of depression-associated tweets in each of six topics (tweets assigned to their dominant topic). Bubble size shows the number of tweets; tick labels give each topic's three highest-weighted words; the dashed line marks the corpus average (52.0%). The analysis used only words written by at least five different users.

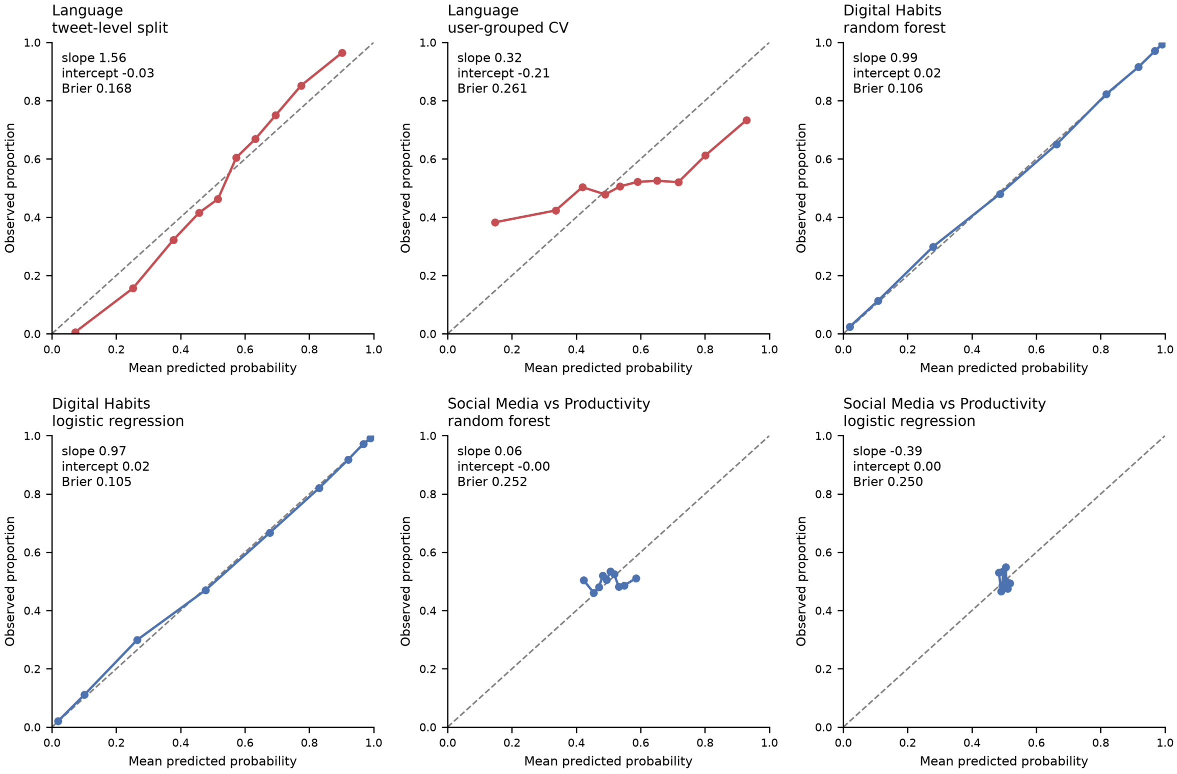


**Figure S4: Calibration of the primary models.** Observed proportion of the positive class against mean predicted probability in deciles of prediction, with calibration slope, calibration-in-the-large (intercept) and Brier score. The dashed line marks perfect calibration. Language models: depression-associated account; behavioural models: higher stress (score $\geq 6$).

**Table S2: Subgroup performance of the random forest in the Social Media vs Productivity dataset.**

| Subgroup | Test records | AUC (95% CI) | Balanced accuracy |
|---|---|---|---|
| All test records | 5,620 | 0.506 (0.490–0.521) | 0.506 |
| *Gender* | | | |
| Female | 2,680 | 0.504 (0.483–0.525) | 0.500 |
| Male | 2,715 | 0.502 (0.482–0.523) | 0.508 |
| Other | 225 | 0.564 (0.493–0.644) | 0.552 |
| *Job type* | | | |
| Education | 987 | 0.497 (0.461–0.531) | 0.498 |
| Finance | 922 | 0.503 (0.466–0.539) | 0.497 |
| Health | 932 | 0.517 (0.477–0.554) | 0.525 |
| IT | 966 | 0.512 (0.476–0.547) | 0.510 |
| Student | 946 | 0.499 (0.466–0.538) | 0.501 |
| Unemployed | 867 | 0.504 (0.465–0.543) | 0.504 |

Primary random forest (stress $\geq 6$) evaluated on the held-out test set. Confidence intervals from 500 bootstrap resamples within each subgroup. Performance is at chance overall and in every subgroup, so the dataset offers no discriminative signal whose fairness could be audited.
